# Benchmarking Large Language Models and Clinicians Using Locally Generated Primary Healthcare Vignettes in Kenya

**DOI:** 10.1101/2025.10.25.25338798

**Authors:** Paul Mwaniki, Wilkister Musau, Lynda Isaaka, Conrad Wanyama, Vaishnavi Menon, Alastair Denniston, Xiaoxuan Liu, Mira Emmanuel-Fabula, Gwydion Williams, Bilal A. Mateen, Ambrose Agweyu

## Abstract

**Background:** Large language models (LLMs) show promise on healthcare tasks, yet most evaluations emphasize multiple-choice accuracy rather than open-ended reasoning. Evidence from low-resource settings remains limited.

**Methods:** We benchmarked five LLMs (GPT-4.1, Gemini-2.5-Flash, DeepSeek-R1, MedGemma, and o3) against Kenyan clinicians, using a randomly subsampled dataset of 507 vignettes (from a larger pool of 5,107 clinical scenarios) spanning 12 nursing competency categories. Blinded physician panels rated responses using a 5- point Likert scale on an 11-domain rubric covering accuracy, safety, contextual appropriateness, and communication. We summarized mean scores and used Bayesian ordinal logistic regression to estimate probabilities of high-quality ratings (≥4) and to perform pairwise comparisons between LLMs and clinicians.

**Findings:** Clinician mean ratings were lower than those for LLMs in 9/11 domains: 2.86 vs 4.25-4.72 (guideline alignment), 2.76 vs 4.25-4.73 (expert knowledge), 2.96 vs 4.30-4.73 (logical coherence), and 2.58 vs 4.16-4.68 (low omission of critical information). On safety-related domains, LLMs received higher ratings: minimal extent of possible harm 3.16 vs 4.29-4.68; low likelihood of harm 3.68 vs 4.54-4.81. Performance was similar for low inclusion of irrelevant content (4.28 vs 4.25-4.35) and for avoidance of demographic bias (4.86 vs 4.91-4.94). In Bayesian models, LLMs had >90% probability of ratings ≥4 in most domains, whereas clinicians exceeded 90% only for contextual relevance and demographic/socio-economic bias. Pairwise contrasts showed broadly overlapping credible intervals among LLMs, with o3 leading numerically most domains except contextual relevance, demographic/socio-economic bias, and relevance to the question. Generating all LLM responses cost USD 3.86–8.68 per model (USD 0.008-0.017 per vignette), compared with USD 3.35 per clinician-generated vignette.

**Interpretation:** LLMs produced responses that were more accurate, safer, and more structured than clinicians in vignette-based tasks. Findings support further evaluation of LLMs as decision support in resource-constrained health systems.

**Funding Statement:** This study was supported by the Gates Foundation [INV-068056].

**Research in Context:** *Evidence before this study:* We searched PubMed, medRxiv, and arXiv (Jan 1, 2021–Sept 30, 2025) using combinations of terms including “large language model”, “LLM”, “healthcare”, “benchmarking”, “clinical decision support”, and “low-resource settings”. The search returned 28 preprints and only 4 peer-reviewed articles. A study from Rwanda benchmarked five LLMs against clinicians using 524 real-world questions from community health workers; all models outperformed clinicians, including in Kinyarwanda (Rutunda, 2025). In Kenya, a multimodal LLM (POE) outperformed primary care providers on 63 otolaryngology cases (79.4% vs 50.8%) and aligned with specialist recommendations (Lechien, 2025). A cross-country maternal health study evaluated GPT-4, GPT-3.5, a custom GPT-3.5, and Meditron-70b on three questions, with expert reviewers in Brazil, Pakistan, and the USA rating outputs in their native languages. GPT-4 and GPT-3.5 were most accurate, though readability and gender bias were noted (Lima, 2025). AraSum, a lightweight Arabic summarization model, outperformed the Arabic foundation model JAIS-30B on BLEU, ROUGE, and expert ratings of accuracy, comprehensiveness, and clinical utility (Lee, 2025). Additional preprints proposed expert-rated benchmarks for LMIC clinical tasks.

*Added value of this study:* This study uniquely combines local co-design, real-world clinical scenarios, and structured, expert-based assessment across 11 dimensions of clinical quality. It demonstrates the relative strengths and weaknesses of five widely available LLMs versus frontline clinician performance, offering evidence of systematic clinician gaps in accuracy, guideline adherence, and completeness.

*Implications of all the available evidence:* LLMs show substantial promise as clinical decision support tools in low-resource health systems. Across multiple settings and task types, current models consistently meet or exceed clinician performance in controlled evaluations. However, real-world deployment requires attention to equity, local clinical validation, and thoughtful implementation pathways that mitigate risk and reinforce trust.

## Introduction

Large language models (LLMs) have shown impressive capabilities in several health-specific evalution exercises,^1-3^but many of these historical benchmarking tasks have focused narrowly on multiple-choice accuracy, which fail to reflect the complexity of real clinical decision-making.^4^ LLMs that excel in multiple-choice exams have been found to struggle with tasks like gathering the right information, following clinical guidelines, and integrating into realistic clinical workflows.^5,6^This highlights the need to design evaluations that incorporate uncertain information, open-ended reasoning, and context-specific criteria.^4,7^ These concerns have prompted a new wave of physician-designed, rubric-based benchmarks that have moved beyond simple question-and-answer toward multidimensional assessments of open-ended performance. Examples include OpenAI’s HealthBench, which evaluates model responses in realistic medical conversations using a detailed rubric with thousands of physician-crafted criteria.^8^ Models are scored along axes such as factual accuracy, completeness, clarity of explanation, contextual awareness, communication quality, and instruction-following. Similarly, the MultiMedQA framework introduced by Google employs expert human raters (both physicians and laypeople) to judge answers on multiple criteria, including clinical correctness, possible harm, and empathy.^9^

In the field of Global Health, there are additional nuances to consider. First, linguistic and cultural under-representation persists in current models. State-of-the-art LLMs trained predominantly on biomedical text from native English-speaking contexts perform worse on African languages and other languages from low-resource settings.^10^ They often struggle with culturally specific content or local context that was absent from their training data. Encouragingly, new research is beginning to close these gaps: one recent study created a million-word benchmark across eight low-resource African languages and demonstrated that targeted fine-tuning, cross-lingual transfer learning, and culturally relevant adjustments can significantly improve an LLM’s performance in those languages.^11^ Nonetheless, the disparities remain an urgent equity concern with current LLMs performing least reliably for populations who have the most to gain from them. Second, tasks must reflect the realities of frontline clinical practice in low- and middle-income country (LMIC) health systems (e.g., where patients often present with incomplete or ambiguous information, and resource constraints often prevent comprehensive diagnostic testing), if they are to be relevant. For example, whilst a treatment recommendation might be correct in principle, if the suggested drug (or intervention more broadly) is unavailable in a rural clinic, the advice is not relevant. There remains a paucity of robust, vignette-style benchmarks generated by frontline healthcare workers in LMICs.

To address this gap, we undertook a benchmarking study of five LLMs side-by-side with practicing Kenyan clinicians (nurses and clinical officers) on a dataset comprising real-world scenarios co-designed and written by Kenyan nurses to reflect authentic patient presentations and challenges in local clinics. We sought to address two broad questions: (1) Can globally trained LLMs generalize to the realities of frontline practice in Kenya? (2) How do these models perform relative to human clinicians when judged on clinical appropriateness, safety, and communication?

## Methods

### Study Design

We conducted a cross-sectional benchmarking study to evaluate the performance of large language models (LLMs) relative to Kenyan clinicians using a dataset of primary care scenarios derived from frontline nursing practice.

### Dataset

The source benchmark dataset consisted of 7,606 clinical scenarios co-designed and generated by 145 nurses across three counties in Kenya: Kiambu, Kakamega, and Uasin Gishu. These scenarios were produced using an adapted Situation-Background-Assessment-Recommendation (SBAR) framework during participatory workshops and follow-up digital submissions.^12^ Nurses contributed scenarios that captured both clinical and non-clinical decision points, reflecting the realities of primary care, including resource limitations, referral dilemmas, and counselling challenges.

Clinicians that contributed responses to the vignettes were selected for their primary care experience and clinical engagement. All 51 were medical graduates. The majority were male (34/51, 66.7%), and the rest were female. Most had less than one year of professional experience (27/51, 52.9%), with 9 (17.6%) reporting between one and seven years, and one clinician having over 20 years of experience (sTable 1). The process of generating clinician responses began with a four-day workshop where participants independently answered 11 randomly selected vignettes, then reached consensus on responses to standardize their approach. Subsequently, the complete pool of clinicians was assigned the remaining vignettes. All responses were reviewed against predefined quality control criteria, and 231 low-quality cases were excluded. Hand-written answers were then compiled and transcribed, yielding responses for 5,107 vignettes.

### Sampling

A stratified random sample of 507 vignettes was drawn from the complete pool. Vignettes were stratified into twelve nursing competency categories: Adult Health, Child Health, General Emergency, Maternal and Child Health, Paediatric Emergency Care, Surgical Care, Mental Health, Neonatal Care, Critical Care, Gender-Based Violence Emergency Care, Sexual and Reproductive Health, and Palliative Care. From each stratum, up to 46 vignettes were randomly selected. Where a category contained fewer than 46 vignettes, all available scenarios were included. This ensured broad coverage of the most common and high-stakes decision points encountered in frontline care.

### Large Language Models

We evaluated five LLMs that were publicly available through APIs or hosted platforms at the time of the study. These included GPT-4.1 (OpenAI), Gemini-2.5-Flash (Google), DeepSeek-R1 (DeepSeek), MedGemma (Google), and o3 (OpenAI). All models were accessed between 21st July 2025 and 4th August 2025, with outputs generated in English to align with the language of nursing education, clinical documentation, and guideline dissemination in Kenya. Responses from LLMs were generated through API calls implemented in Python using the LangChain library. Access to DeepSeek-R1 and MedGemma was provided through Hugging Face, Gemini-2.5-Flash through Google AI Studio, and GPT-4.1 and o3 through OpenAI. Model prompts were standardised across platforms to ensure consistency, with instructions to provide clinical reasoning, recommendations, and any relevant contextual considerations (See Box 1).

#### Box 1

*LLM Prompt*

*“You (the LLM) are a professor of primary healthcare in Kenya. You will be receiving queries from nurses as part of an experiment, and you should answer the questions to the best of your abilities, and use language and guidance that is contextually appropriate and relevant to the practice of medicine by a community nurse in Kenya. Leverage local guidelines where appropriate to inform your responses. The next thing you receive will be the nurse’s communication*.*”*

### Evaluation Framework

Responses from both clinicians and models were evaluated using an 11-domain rubric adapted from existing frameworks for medical AI evaluation.^13^ The domains comprised: (1) alignment with established medical guidelines and evidence; (2) accuracy and expert-level knowledge; (3) clarity and professionalism of presentation; (4) logical structuring and coherence; (5) consideration of cultural, regional, and resource-specific factors; (6) avoidance of demographic bias; (7) likelihood of harm if followed; (8) severity of potential harm if followed; (9) omission of critical information; (10) inclusion of unnecessary information; and (11) accurate understanding and addressing of the question. Each domain was scored on a five-point Likert scale ranging from 1 (very poor) to 5 (excellent). Evaluators were instructed to consider both clinical content and communicative quality when assigning scores.

### Expert Panel

A panel of six family physicians conducted the evaluation; five female and one male, each with 10 -16 years of ongoing professional experience practicing in Kenya. They independently scored responses to all vignettes in pairs within their assigned sets, which included both clinician and model-generated outputs. To minimize bias, the origin of each response was concealed from the evaluators. Discrepancies of greater than one point between raters were reviewed in consensus meetings. If consensus could not be reached, a third panel member adjudicated the final score. When scores differed by a single point, the lower score was conservatively retained. Panel evaluations were conducted between 4th August 2025 and 29th August 2025.

### Statistical Analysis

Descriptive statistics were computed as means and standard deviations for each evaluation domain by respondent type (clinician or LLM). We also estimated the costs of generating responses from clinicians and for each model, with per-vignette estimates calculated where feasible to facilitate comparability.

To formally compare ratings, Bayesian ordinal logistic regression models were fitted using the brms package in R (version 4.5.1).^14^The outcome was the domain-specific rating, and predictors included the response source (clinician or LLM). Models incorporated varying intercepts for panel and fixed effect interactions between response source and domain. Broad priors were specified. Four Markov Chain Monte Carlo (MCMC) chains were run, each with 1,000 warm-up iterations and 4,000 post-warm-up draws, yielding a total of 16,000 post-warm-up samples. Convergence was assessed using 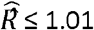 and effective sample size diagnostics, and posterior predictive checks indicated good model fit. Model specification and fitting details are available in sBox 1.

Posterior draws from these models were summarized to estimate domain-specific probabilities of achieving high-quality ratings (≥ 4), and pairwise contrasts between response sources expressed as differences in log-odds. These complementary analyses allowed both absolute and relative comparisons of clinician and LLM performance while quantifying uncertainty through posterior credible intervals. Analysis scripts are available in our GitHub repository and archived on Zenodo.^15^

### Ethical Considerations

Ethical approval for the dataset development and benchmarking study was obtained from the Maseno University Institutional Review Board (Ref: MSU/DRPI/MUERC/00899/20). All participating nurses and clinicians provided written informed consent. Data were de-identified prior to analysis to protect confidentiality.

## Results

All nursing competency categories contributed 46 vignettes each except critical care (n=29) and palliative care (n=18). Fifteen clinician responses were excluded from expert panel evaluation because they were found to be inconsistent with the nurses’ questions. Figure 1 summarizes the data generation and evaluation process, and the characteristics of the clinicians that contributed responses are summarized in sTable 1.

**Figure 1.**
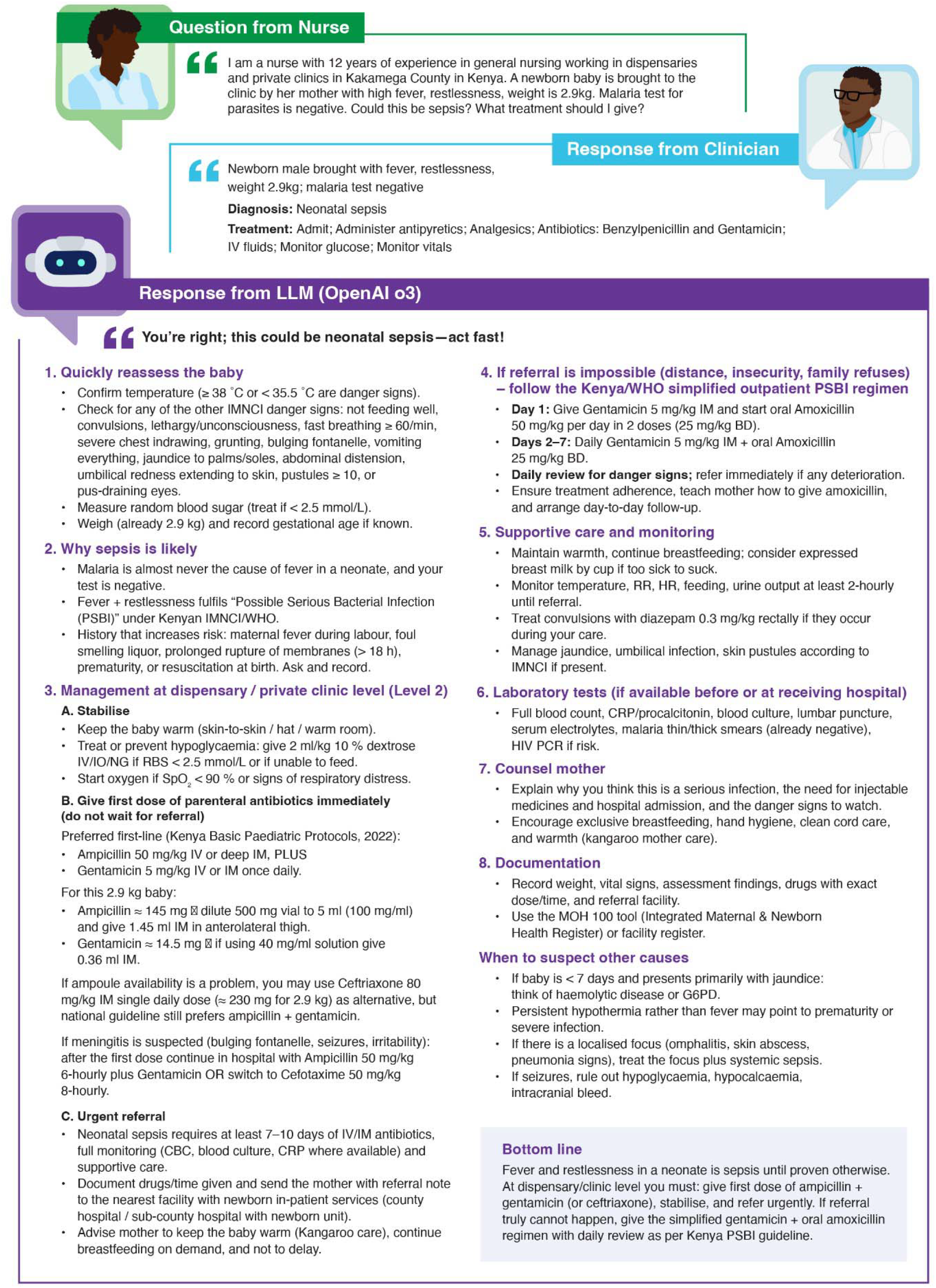
Data Generation and Evaluation Process

### Clinician Scored Lower Than LLMs Across Most Domains

The largest gaps were observed in alignment with medical guidelines, expert-level knowledge base, logical consistency, omission of critical information, and severity of harm. Across all domains, the LLMs clustered closely together. See Figure 2 for a visual summary of comparative performance across the 11 evaluation domains, and sTable 2 for the full numerical summary.

**Figure 2.**
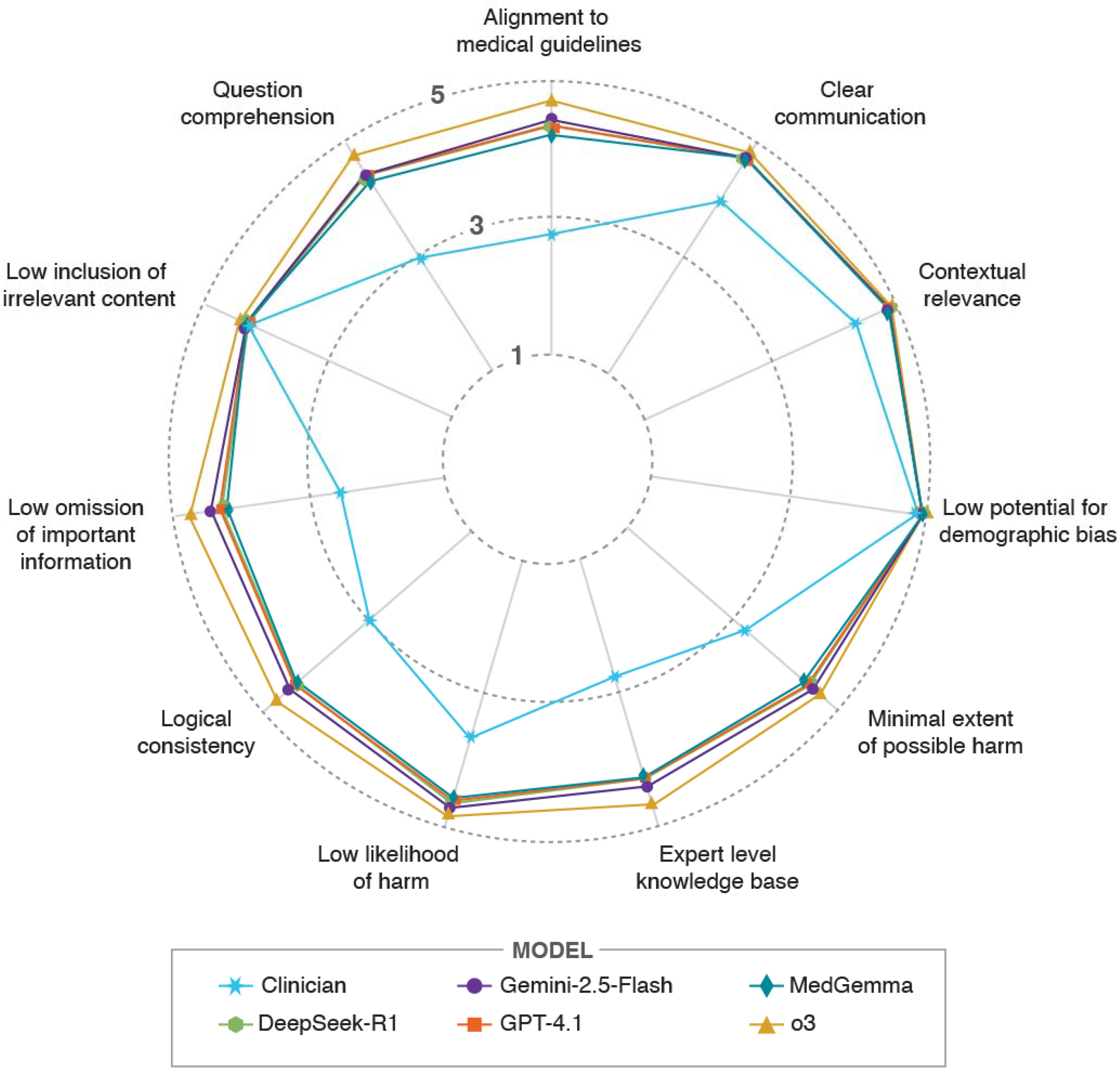
Radar plot of clinician and LLM performance profiles across 11 evaluation domains

For alignment with established medical guidelines, clinicians achieved a mean rating of 2.86 (SD 0.72), compared with 4.25-4.72 for LLMs, with o3 performing best. Clarity of responses was also lower for clinicians (3.95, SD 0.90) relative to the consistently high ratings for LLMs (4.65-4.80). Consideration of regional, cultural, and resource-specific factors was strong in both groups, but more consistent among LLMs, with o3 again rated highest (4.86, SD 0.39 vs 4.33, SD 0.93 for clinicians). Avoidance of demographic bias approached the maximum across all models, with minimal difference between clinicians (4.86) and LLMs (4.91-4.94).

Large disparities were observed in safety-related domains. Clinicians scored 3.16 (SD 1.07) for minimal extent of possible harm and 2.76 (SD 0.67) for accuracy and expert-level knowledge, whereas LLMs scored above 4.25 in both domains, with o3 again highest. Low likelihood of harm if advice was followed was also lower for clinicians (3.68, SD 1.01) than for all LLMs (≥ 4.54). Logical structuring and coherence was another area of weakness for clinicians (2.96, SD 0.56), compared with 4.30-4.73 for LLMs.

Clinicians were more likely to omit critical information (2.58, SD 0.87), while most LLMs reduced such omissions and o3 achieved the highest rating (4.68, SD 0.51). In contrast, the inclusion of unnecessary information was similar across groups, with little difference between clinicians (4.28) and LLMs (4.25-4.35). Finally, the ability to understand and address the question asked was rated substantially lower for clinicians (2.99, SD 0.61) than for LLMs (≥ 4.32), with o3 again the top performer (4.74, SD 0.45) (sTable 1).

### Persistent Clinician–LLM Gaps and Comparable Performance among LLMs in Adjusted Analyses

Bayesian models estimating the probability of responses being rated as high quality (≥ 4) revealed marked differences between clinicians and LLMs across the 11 evaluation domains (sFigure 1). For clinicians, the estimated probability of achieving a high-quality rating was below 0.5 in several domains, including alignment with medical guidelines, expert-level knowledge base, logical consistency, severity of harm, and understanding of the question. The lowest probability was observed for omission of critical information, where clinicians had only a 11% chance of reaching the high-quality threshold. By contrast, all five LLMs demonstrated probabilities consistently above 0.9 across all domains, with overlapping credible intervals indicating similar performance, and o3 generally achieving the highest estimates. Domains such as demographic and socio-economic bias and contextual relevance showed near-ceiling performance for both clinicians and LLMs, suggesting little room for differentiation. Clinicians’ performance matched the LLMs for demographic and socio-economic bias, as indicated by overlapping credible intervals. Smaller but still notable gaps were evident for clear communication and relevance to the question, where clinicians performed better than in other domains yet remained below the consistently high LLM estimates.

We undertook pairwise comparisons of models across the 11 evaluation domains (Figure 3). Across nearly all domains, each LLM significantly outperformed clinicians, with differences most pronounced for alignment with medical guidelines, expert-level knowledge base, logical consistency, severity of harm, and understanding of the question. Clinician disadvantage was particularly severe in the omission of critical information, where all LLMs demonstrated substantially higher log-odds of achieving high-quality ratings. In contrast, differences between clinicians and LLMs narrowed in demographic and socio-economic bias avoidance, and relevance to the question, where difference in log odds clustered close to zero. Comparisons among the LLMs themselves revealed overlapping credible intervals in most domains, indicating broadly similar performance, though o3 tended to outperform other models across all domains, except in contextual relevance, demographic and socio-economic bias and relevance to the question.

**Figure 3.**
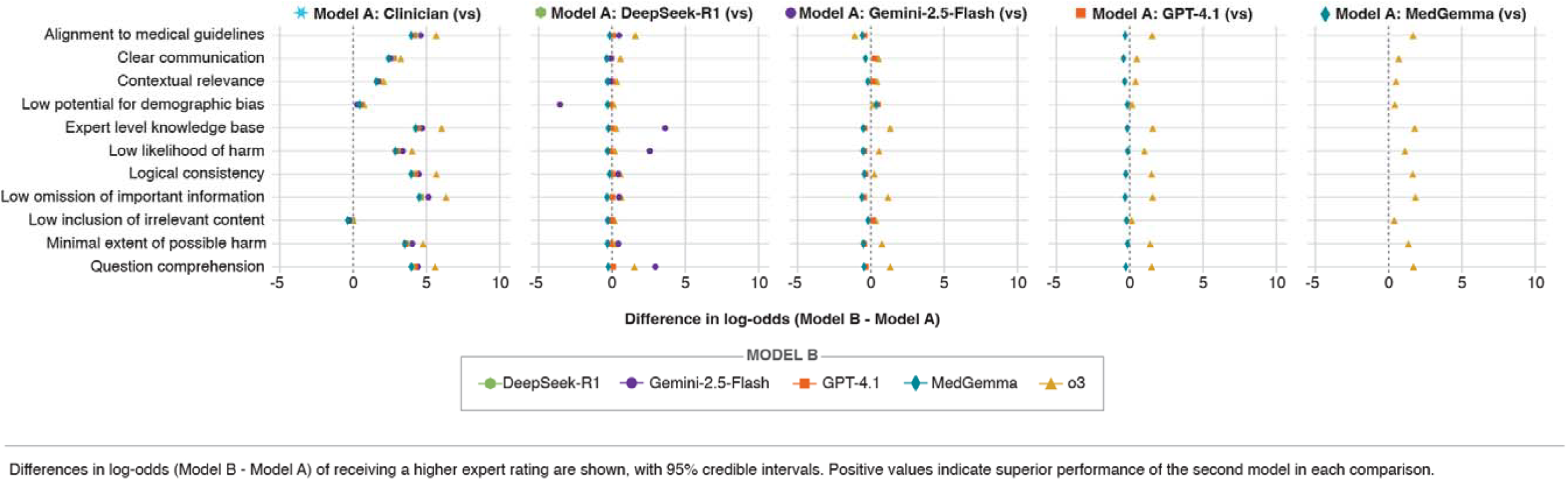
Pairwise comparisons of clinicians and LLMs across 11 evaluation domains.

### GenAI-generated Responses Are At Least 100x Cheaper Than Human Responses

The cost of generating responses varied modestly across models. For the 507 vignettes, the total cost was USD 4.26 for Gemini-2.5-Flash, USD 3.86 for GPT-4.1, and USD 8.68 for o3, corresponding to unit costs of approximately USD 0.008, USD 0.008, and USD 0.017 per vignette, respectively. Itemized costs could not be estimated directly for DeepSeek-R1 and MedGemma: access to DeepSeek-R1 was covered by a flat USD 9 monthly subscription, while MedGemma required deployment of an inference endpoint billed at USD 2.50 per hour. By comparison, for the 7,606 clinical scenarios generated overall, the total cost was USD 25,077 (USD 492 per clinician for 51 clinicians), corresponding to an average cost of about USD 3.35 per vignette.

## Discussion

In this benchmarking study of clinical vignettes spanning multiple nursing competency categories, LLMs outperformed clinicians across most response-quality domains. The greatest differences were observed in safety-related measures and the accuracy of clinical reasoning. Clinicians frequently omitted critical information, demonstrated weaker logical structure, and scored lower for accuracy and expert-level knowledge, whereas LLMs produced responses that were more complete, coherent, and closely aligned with clinical guidelines.

o3 achieved the highest numerical mean scores across nearly every domain, including those often regarded as challenging for automated systems such as contextual sensitivity to regional, cultural, and resource-specific factors.^16,17^Gemini-2.5-Flash, GPT-4.1, DeepSeek-R1, and MedGemma also performed substantially better than clinicians, with the probability of a high rating being higher across all domains except demographic and socio-economic bias. Pairwise comparisons between LLMs showed broadly overlapping credible intervals, indicating similar levels of performance, although o3 tended to outperform the others in knowledge- and safety-related domains.

Performance was broadly comparable between clinicians and LLMs in avoiding unnecessary information, and all responders scored near the ceiling for avoidance of demographic bias. Across the remaining domains, LLMs consistently surpassed human performance, providing strong evidence that contemporary models can generate responses that are not only clinically accurate but also safer, more structured, and more contextually appropriate than those of clinicians.

### Results in Context of the Literature

Our findings align with and extend the evidence base on LLMs in healthcare from other LMICs. A recent benchmarking study from Rwanda compared frontier LLMs with local clinicians using locally developed vignettes and found model advantages of +0.7-0.9 points (relative to human clinicians) on a 5-point scale across 11 expert-rated domains. Although performance dipped modestly (0.2-0.3 points) when vignettes were administered in Kinyarwanda, LLMs still outperformed clinicians overall.^18^Taken together with our Kenyan benchmark, these results provide cross-country evidence that globally trained models generalize effectively to frontline African health care, while underscoring the need for localisation strategies to address language and contextual gaps.

Our results differ from earlier evaluations that highlighted LLMs’ susceptibility to hallucinations and unsafe recommendations.^19,20^Although hallucinations were not directly measured in this study, our rubric captured related safety dimensions, such as harmful recommendations and omissions where newer-generation models, particularly o3, performed strongly. This suggests that the pace of model development is addressing some of the shortcomings identified in earlier generations.

The findings of this study contribute to conversations relating to expanding clinical use-cases of AI in healthcare.^19,21,22^Recent reviews emphasize that rigorous evaluation is needed to ensure the reliability and safety of LLMs before integrating them into complex real-world clinical contexts.^23^The superior performance of LLMs over clinicians across multiple domains of quality suggests that these tools could play a role, particularly in environments where frontline health workers often face high patient loads, limited supervision, and constrained access to specialist expertise. The strong performance of LLMs in domains directly linked to patient safety is particularly compelling in this regard. Although our study was designed as a head-to-head comparison between clinicians and LLMs, these findings should not be interpreted as support for replacement. The benchmarking design enabled us to quantify relative strengths and weaknesses. In practice, the most promising role for LLMs is likely to complement and extend human expertise, with clinicians retaining oversight.^6,24^

### Implications for Policy Makers

From a policy perspective, these results underscore the urgency of strengthening (and, where absent, developing) regulatory and governance frameworks that can both enable innovation (i.e., leveraging the potential of these tools’ supra-human capabilities), whilst safeguarding against risks.^25^For example, most of the models evaluated in this study are closed-weight, API-based systems that transmit data to external servers for inference. This architecture raises concerns about patient data confidentiality, compliance with local data protection frameworks, and the ability to conduct audits or maintain on-premises control over sensitive clinical information. DeepSeek-R1 and MedGemma are open-weight, meaning their parameters are publicly available and can be deployed in secure, self-hosted environments. MedGemma, being relatively lightweight, can potentially run locally on modest hardware, offering a more privacy-preserving alternative for clinical or research settings. In short, there is more to consider than raw performance when selecting the most appropriate LLM for a given task, and many countries lack guidance on how to effectively manage these trade-offs.

At the same time, integrating LLMs into practice raises important questions for workforce development. If LLMs become trusted sources of clinical guidance, there is a risk that health workers may over-rely on these systems, potentially leading to deskilling.^26^Conversely, if deployed thoughtfully, LLMs could serve as powerful educational tools, providing frontline workers with exposure to expert reasoning patterns and up-to-date guideline-based recommendations in real time. Whether these systems ultimately augment or erode human expertise will depend on how they are embedded into health systems and the safeguards put in place.

Finally, the cost analysis and economic implications are also notable. Generating responses for 507 vignettes cost less than USD 10 for most models, corresponding to per-vignette costs of approximately USD 0.008 for Gemini-2.5-Flash and GPT-4.1, and USD 0.017 for o3. This contrasts with approximately USD 3 per vignette for the clinician-generated responses. Relative to the costs of conventional training or continuous professional development programmes, the marginal cost of integrating LLMs into clinical workflows appears minimal. Although true deployment costs will depend on infrastructure, access models, and ongoing maintenance, these findings suggest that inference-specific resource requirements alone are unlikely to be a barrier to LLM deployment. For policymakers in LMICs, this represents a unique opportunity to overcome traditional barriers to scaling expertise, provided that investments are made in safe and equitable implementation.

### Strengths & Limitations

This study represents one of the few comprehensive benchmarking exercises in low resource settings. The breadth of scenarios allowed us to assess performance across diverse aspects of clinical reasoning, including guideline adherence, factual accuracy, patient safety, contextual sensitivity, and communication quality. The vignette corpus was locally co-designed, enhancing contextual relevance. By grounding the cases in the realities of frontline nursing and primary care practice, the evaluation provides a closer approximation of the challenges faced by health workers in low- and middle-income settings, where the potential impact of LLM-based decision support is greatest. Moreover, we applied Bayesian ordinal regression, which offered several methodological advantages: quantification of uncertainty, and direct estimation of posterior probabilities of differences between clinicians and LLMs.^27^This approach avoided repeated pairwise testing and provided interpretable, probabilistic statements about model performance.

We also acknowledge several limitations. The study relied on vignette-based assessments rather than real patient encounters. Although vignettes are a widely used and validated method for benchmarking decision support systems, they cannot replicate the full complexity of clinical practice, which includes iterative history-taking, physical examination, patient communication, non-verbal cues, and decision-making under time pressure.^4^As such, our findings may over- or under-estimate performance in real-world settings. Notably, this evaluation was limited to single-turn prompts and responses, whereas the prevailing belief is that we need to better reflect the process of clinical reasoning, which often unfolds over multi-turn interactions.^28^That said, evidence is beginning to emerge that even single-turn interactions can be useful: a retrospective analysis we conducted using routine clinical records confirmed the promise of LLM-based decision support in practice,^29^and a multicentre pragmatic trial currently underway is testing these systems prospectively.^30^Furthermore, using nurses to generate the vignettes, because of the breadth of their primary care responsibilities, runs the risk of not fully representing the experiences of all community-based health workers at this level, particularly community health workers, who are often the intended users of decision support systems in low-resource settings.^31^Finally, the language of evaluation was restricted to English. Although our rubric assessed contextual sensitivity, we did not benchmark performance in Swahili or other local languages widely used in Kenyan healthcare. In reality, patient-provider exchanges are complex, often involving a mix of languages, dialects, and code-switching that vary across geographies and patient populations.^32^Evidence from Rwanda suggests that model performance dips modestly when tasks are presented in local languages rather than English ^18^. Addressing these gaps will be critical for equitable deployment in multilingual health systems.

## Conclusions

This study provides robust evidence that contemporary LLMs can outperform clinicians in generating high-quality responses to clinical vignettes across a broad spectrum of competency domains. The most striking differences were observed in safety-related measures and accuracy of reasoning, with o3 demonstrating the most consistent gains. These advantages extended beyond factual correctness to include contextual sensitivity and logical structuring, attributes central to safe clinical care.

Our findings suggest that LLMs have the potential to serve as powerful clinical decision support tools, particularly in low resource settings where access to expertise is constrained. Their low marginal cost further supports feasibility. Realising this promise, however, will require rigorous real-world evaluation, regulatory safeguards, and integration strategies that prioritize equity and workforce development. If these conditions are met, LLMs could play a transformative role in strengthening the quality, accessibility, and safety of healthcare delivery worldwide.

## Data Availability Statement

De-identified quantitative data and all analysis code supporting the findings of this study are openly available at: https://github.com/pmwaniki/vignette/tree/v1.0.1, and the associated Zenodo archive (10.5281/zenodo.17340120.). The repository includes datasets, analysis scripts, and documentation sufficient to reproduce the primary results and figures.

## Patient and Public Involvement Statement

Patients were not involved as research participants.

## Author contributions (CRediT taxonomy)

Conceptualization: BAM; Methodology: all authors; Data curation (extraction, de-identification, management): WM; Investigation: all authors; Formal analysis: PM; Project administration: WM, LI, CW; Supervision: BAM, AA. Funding acquisition: BAM. Writing (original draft): PM, AA; Writing (review and editing): all authors. AA is the study’s guarantor.

## Acknowledgments

We thank the experts who served on the evaluation panel: Dr Kaya Belknap, Dr Gad Igiraneza, Dr Faith Lelei, Dr Miriam Miima, Dr Liliane Mugodo, and Dr Sheila Sisela.

