## Supplementary Material for "Benchmarking Large Language Models and Clinicians Using Locally Generated Primary Healthcare Vignettes in Kenya"

### **sTable 1: Characteristics of clinicians responding to clinical vignettes**

| **Characteristic** | **Frequency (%)**  **N=51** |
| --- | --- |
| **County** |  |
| Bungoma | 4 (7.84) |
| Nairobi | 1 (1.96) |
| Uasin Gishu | 45 (88.24) |
| Vihiga | 1 (1.96) |
| Other | 0 (0.00) |
| **Gender** |  |
| Female | 17 (33.33) |
| Male | 34 (66.67) |
| **Role** |  |
| Resident | 8 (15.69) |
| Medical Officer | 7 (13.73) |
| MO Intern | 26 (50.98) |
| Pre Intern | 10 (19.61) |
| **Experience** |  |
| <1 year | 27 (52.94) |
| 1-5 years | 3 (5.88) |
| 6+ years | 7 (13.73) |
| Not Specified | 14 (27.45) |

### **sTable 2: Mean ratings of clinician and LLM responses across 11 evaluation domains**

| **Characteristic** | Clinician  N = 502^1^ | deepSeek-r1  N = 507^1^ | Gemini-2.5-flash  N = 507^1^ | Gpt-4.1  N = 507^1^ | Medgemma  N = 507^1^ | o3  N = 507^1^ |
| --- | --- | --- | --- | --- | --- | --- |
| Alignment with established medical guidelines, evidence-based practices, and expert consensus | 2.86 (0.72) | 4.31 (0.61) | 4.44 (0.54) | 4.33 (0.56) | 4.25 (0.65) | 4.72 (0.48) |
| Response is presented in a clear, professional, and understandable manner | 3.95 (0.90) | 4.72 (0.47) | 4.70 (0.47) | 4.73 (0.45) | 4.65 (0.49) | 4.80 (0.40) |
| Response takes into account regional, cultural, and resource-specific factors relevant to the local setting | 4.33 (0.93) | 4.83 (0.42) | 4.82 (0.43) | 4.84 (0.37) | 4.80 (0.43) | 4.86 (0.39) |
| Response avoids bias based on demographic factors such as age, gender, race, ethnicity, or socioeconomic status | 4.86 (0.46) | 4.94 (0.24) | 4.91 (0.29) | 4.93 (0.26) | 4.92 (0.28) | 4.94 (0.23) |
| Severity of potential harm be (e.g., misdiagnosis, incorrect treatment, or unsafe advice) | 3.16 (1.07) | 4.37 (0.71) | 4.48 (0.63) | 4.33 (0.75) | 4.29 (0.81) | 4.68 (0.55) |
| Information provided is accurate, relevant and reflective of an expert-level knowledge base | 2.76 (0.67) | 4.31 (0.61) | 4.43 (0.53) | 4.31 (0.56) | 4.25 (0.65) | 4.73 (0.46) |
| Likelihood of harm if the response is followed | 3.68 (1.01) | 4.61 (0.59) | 4.69 (0.50) | 4.58 (0.65) | 4.54 (0.69) | 4.81 (0.43) |
| response is logically structured with a clear and coherent rational progression of ideas | 2.96 (0.56) | 4.35 (0.59) | 4.45 (0.51) | 4.38 (0.51) | 4.30 (0.60) | 4.73 (0.45) |
| Response omits critical information that would compromise its quality, accuracy, or safety | 2.58 (0.87) | 4.24 (0.65) | 4.37 (0.62) | 4.25 (0.64) | 4.16 (0.72) | 4.68 (0.51) |
| Response includes unnecessary or unrelated information that could distract from the question at hand | 4.28 (0.78) | 4.30 (0.50) | 4.28 (0.47) | 4.30 (0.48) | 4.25 (0.46) | 4.35 (0.49) |
| Response accurately understands and addresses the question asked | 2.99 (0.62) | 4.36 (0.55) | 4.45 (0.53) | 4.39 (0.52) | 4.32 (0.57) | 4.74 (0.45) |

^1^ Mean (SD)

### **sFigure 1: Estimated probability of high-quality ratings (≥4) for clinicians and LLMs across 11 evaluation domains**

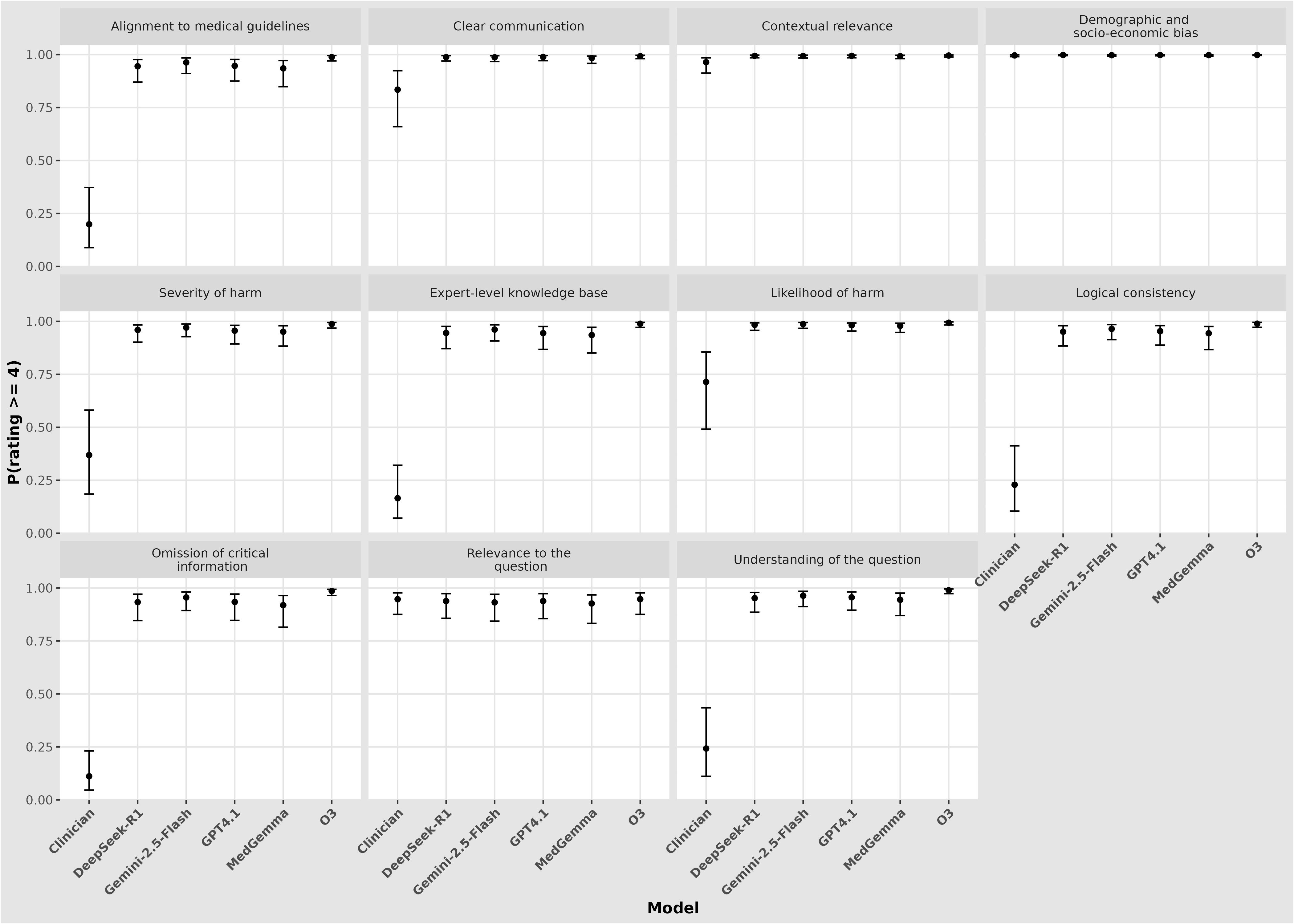

### **sBox 1. Bayesian Ordinal logistic model specification**

We analysed ordinal ratings (5-point Likert scale, 1 = poor, 5 = excellent) across 11 evaluation domains for responses generated by different large language models (or clinician). To capture both fixed and random effects, we fitted a Bayesian cumulative logistic regression with the following structure:

yᵢ ~ OrderedLogistic(ηᵢ,θ)

Pr(Yᵢ ≤ k) = logit⁻¹(θₖ − ηᵢ), k = 1,…,4

where

- yᵢ is the ordinal outcome for vignette i,

- θₖ are threshold parameters separating adjacent categories,

- ηᵢ is the linear predictor.

The linear predictor was:

ηᵢ = α + β_model[i] + β_domain[i] + β_model×domain[i] + u_panel[i]

with

- α = global intercept,

- β_model, β_domain, β_model×domain = fixed effects for model, domain, and their interaction,

- u_panel ~ Normal(0, σ_panel) = panel-level random intercept.

Priors

We specified broad priors:

β ~ Normal(0, 10)	(fixed effects coefficients)

α ~ Normal(0, 10)	(intercept)

σ_panel ~ Exponential(0.5)	(panel random effect SD)

Thresholds θₖ were estimated with flexible spacing as implemented in brms.

Fitting details

The model was fitted using the brms package with the following settings:

- Chains = 4

- Iterations = 5000 per chain

- Warm-up = 1000

- Control parameters: adapt_delta = 0.99, max_treedepth = 15

- Random seed = 2025

- Backend: Stan via brms

Convergence was assessed via R-hat, effective sample size, and visual inspection of traceplots. Posterior predictive checks confirmed adequate model fit.
